# Forecasting the Spread of COVID-19 under Different Reopening Strategies

**DOI:** 10.1101/2020.05.26.20113993

**Authors:** Meng Liu, Raphael Thomadsen, Song Yao

**Affiliations:** Olin Business School, Washington University in St. Louis, Missouri, 63130, USA

**Keywords:** COVID-19, SIR models, Social distancing, Reopening

## Abstract

We combine COVID-19 case data with mobility data to estimate a modified susceptible-infected-recovered (SIR) model in the United States. In contrast to a standard SIR model, we find that the incidence of COVID-19 spread is concave in the number of infectious individuals, as would be expected if people have inter-related social networks. This concave shape has a significant impact on forecasted COVID-19 cases. In particular, our model forecasts that the number of COVID-19 cases would only have an exponential growth for a brief period at the beginning of the contagion event or right after a reopening, but would quickly settle into a prolonged period of time with stable, slightly declining levels of disease spread. This pattern is consistent with observed levels of COVID-19 cases in the US, but inconsistent with standard SIR modeling. We forecast rates of new cases for COVID-19 under different social distancing norms and find that if social distancing is eliminated there will be a massive increase in the cases of COVID-19.

## Introduction

We measure the extent to which social distancing reduces the speed at which COVID-19 spreads. We then run simulations to forecast the rates of COVID-19 spread under different social distancing levels. We find that COVID-19 spreads less than proportionately with the number of infectious individuals, a distinct difference from the assumption of standard models. One key implication of this finding is that each additional infectious individual has less impact on the disease spread as more people become infected. We demonstrate that this pattern could be explained by the interconnectedness of people’s social networks. We also observe that social distancing greatly reduces the spread of COVID-19.

The model we estimate is a modified version of a susceptible-infected-recovered (SIR) model. In such models, there is a susceptible population, which is assumed to be equal to the population of whichever region is being examined minus the number of people that have previously had the disease. Some of the susceptible individuals get infected in each period, where the rate of infection is a function of the number of infectious individuals as well as other factors that shift the rate of transmission. Finally, infectious individuals move to a state of recovery. In our analysis, we call anyone who was sick but is no longer infectious to be “recovered,” although some of these people may still actually be sick, hospitalized or have died. Thus, the recovered terminology is actually a shorthand for all post-infectious states.

SIR models and their variants are widely used to characterize infectious disease spread. For example, ^1^ estimates a Susceptible-Exposed-Infected-Confirmed-Removed (SEIQR) model, which adds a stage modeling susceptible people who become exposed to the virus and a stage modeling which infected people are confirmed to have the disease to the standard SIR model. They use this model to look at COVID-19 transmission in Wuhan, China, showing that an earlier lockdown makes the outbreak worse in Wuhan but helps the rest of the world. ^2^ complements ^1^ by using a SEIQR model to show that earlier lockdowns in Wuhan, China reduced the spread of COVID-19 in the Shanxi Province, China. Another paper, ^3^, builds on this insight by showing how transportation seeded the spread of COVID-19 in different Chinese cities. Employing a SEIR model (adding an exposure step to the SIR model) within bats, host animals and humans, ^4^ computes the rate of transmission both from animals to people and from people to people. While it may seem that having more steps in the model would make the SEIR model superior to the SIR model, ^5^ shows that the standard SIR model does a better job at predicting the spread of COVID-19, based on data from Wuhan, China.

Mathematically, we model transmission of COVID-19 as

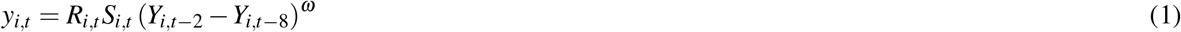

where *y*_*i,t*_ is the number of new infections in county *i* on date *t, R*_*i,t*_ is the rate at which infectious individuals transmit the disease, *S*_*i,t*_ is the percentage of the county population that has not yet had COVID-19, and *Y*_*i,t*_ is the cumulative number of individuals who have been infected by date *t*. Correspondingly, the *Y*_*i,t-*2_ *Y*_*i,t-*8_ term reflects our assumption that infected individuals are infectious from the second day after they catch the virus through the seventh day. This implies that the average serial interval is 4.5 days under the assumption that the level of infectiousness and the level of contact with susceptible individuals is constant during this time ^6^. This treatment of the infectious population is an approximation of the standard SIR model, where the infectious population is typically modeled as a stock that has a constant outflow rate. Discretizing the rate of transmission enables the estimation of a large number of county and date fixed effects in our model, and as a practical matter this assumption has little impact on our estimates of the contagion rate. As a robustness check, we obtain extremely similar COVID-19 forecasts if we take the time of infectiousness to be 14 days, *Y*_*i,t-*2_ *Y*_*i,t-*16_, instead of 6 days, as presented in the appendix. The main difference between our model and the standard SIR model is the inclusion of the exponent *ω* on the number of infectious individuals. This *ω* allows the rate of growth of COVID-19 to be less than proportionate with the number of infectious individuals if *ω*<1. Such a result would be expected if infectious individuals expose many of the same unexposed individuals, which could occur if people have overlapping social connections. We see this directly when, for example, cases are clustered within households, nursing homes, or places of work. Thus, we can think of *ω* as measuring the extent to which people’s networks are more interconnected to a tight-knit group of individuals relative to their level of connectedness to the population as a whole.

We also allow the transmission rate *R*_*i,t*_ to vary with a number of factors instead of treating it as a constant parameter:

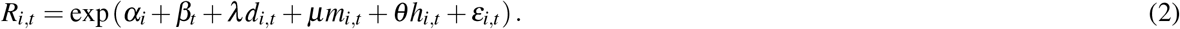

We use *d*_*i,t*_, *m*_*i,t*_, and *h*_*i,t*_ to represent the level of social distancing, temperature, and humidity in county *i* on date *t*, respectively, and *e*_*i,t*_ is the statistical error term. The parameters *α* and *β* are vectors of county and date fixed effects, where the *i*-th element of *α, α*_*i*_, represents the fixed effect for county *i*. Similarly, the *t*-th element of *β, β*_*t*_ represents the fixed effect for date *t*. These fixed effects measure the baseline transmission rate of each county and each date, respectively. The parameters *λ, µ*, and *θ* measure the impacts of social distancing, temperatures, and humidity on transmission rates, respectively. In short, this specification allows transmission rates to differ across counties (through the county fixed effects), dates (through the date fixed effects), levels of social distancing, temperatures, and humidity. We note that the impacts of the last two factors have been debated in the literature^7,8,9^. The county fixed effects account for differences in demographics across counties, such as the demographics shown in Table 2 below as well as other unobservable county-specific factors. The date fixed effects account for both day-of-the-week differences in the patterns of travel for people (e.g., the time away from the house to go to work or to go to the park, which may lead to different exposures to the disease) as well as differences in the rate of testing and reporting that occur across time. As a robustness check, we also include the state-level testing numbers directly into Equation 2 during estimation. The results are statistically indistinguishable from the main results, as noted in **Results and Simulation** below.

**Table 1.** Estimation of a Modified SIR Model.

| Dependent Variable | Log(Infected in County $i$<br>on Date $t$) |
| --- | --- |
| $\lambda$: Impact of Social Dist. Level in<br>County $i$ on Date $t$ | -0.824***<br>(0.245) |
| $\omega$: Impact of Infectious Individuals in<br>County $i$ on Date $t$ | 0.571***<br>(0.014) |
| $\mu$: Impact of Avg. Temperature (Celcius) of<br>County $i$ on Date $t$ | -0.001<br>(0.002) |
| $\theta$: Impact of Avg. Humidity of<br>County $i$ on Date $t$ | 0.005**<br>(0.002) |
| $\alpha$: 2923 County Fixed Effects<br>(Autauga County, AL is omitted as the baseline) | Estimated |
| $\beta$: 101 Date Fixed Effects | Estimated |
| Observations | 131,272 |
| R_squared | 0.63 |
| Counties | 2,924 |

\*\*\* p<0.01, \*\* p<0.05, \* p<0.1

**Table 2.** Analysis of County Fixed Effects.

| Dependent Variable | County Fixed Effect of Each County |
| --- | --- |
| Log(Pop. Density of Each County)<br>(People/Sq. Miles) | 0.4410***<br>(0.0096) |
| Fraction of Black Residents<br>in Each County | 0.8084***<br>(0.0979) |
| Fraction of Hispanic Residents<br>in Each County | 1.3438***<br>(0.1003) |
| Percentage of Commuters using Pub.<br>Transportation in Each County | 5.0215***<br>(0.4140) |
| Log(Median Income of Each County)<br>(in U.S. dollars) | 1.3387***<br>(0.0579) |
| Percentage of Senior<br>Residents of Each County ($\geq 70$yrs) | 1.8825***<br>(0.5255) |
| Percentage of Children<br>Residents of Each County ($< 18$yrs) | 0.6614<br>(0.4733) |
| Constant | -16.6781***<br>(0.6462) |
| R_squared | 0.69 |
| Counties | 2,923 |
| *** p<0.01, ** p<0.05, * p<0.1 |  |

The social distancing measure, *d*_*i,t*_, is based on cellphone GPS location data that are provided by SafeGraph for free to researchers studying COVID-19. We measure social distancing as the first principle component of several daily measures of each county: the percentage of residents staying home, the percentage of residents working at workplace full-time, the percentage of residents working at workplace part-time, the median duration of residents staying home, and the median distance of residents traveled.

As noted earlier, the most crucial difference between our model and a standard SIR model is that a standard SIR model constrains the exponent *ω* = 1. We instead find that *ω* = 0.57. Thus, the marginal impact of one more infected person diminishes as more people are infected. Such a result would be expected if infectious individuals expose many of the same unexposed individuals within a clustered network of individuals. In the appendix we demonstrate that a networking model with contagion can yield *ω <* 1.

## Results and Simulation

The estimated model appears in Table 1, with standard errors (s.e.) reported in the parentheses. The estimated exponent on the number of infectious people, *ω*, is 0.57. Thus, the number of new infections is concave with respect to the number of infectious individuals. This level of concavity also implies that while initial outbreaks of COVID-19 expand exponentially, the daily number of new cases quickly stabilizes to a long-term plateau. We also find that social distancing has a large impact on the growth rate of COVID-19, while humidity has a smaller effect and temperature is insignificant. (When including daily testing numbers of each state in Equation 2, the estimates of *ω* and social distancing are 0.568 (s.e. = 0.014) and −0.816 (s.e. = 0.246), respectively.)

All county-level demographic factors remain constant over time in our analysis. While our main regression gives many insights, impacts of these demographic factors on the spread of the virus are captured by the county fixed effects. In order to better understand how these factors affect the contagion rate, we next regress the county fixed effects *α* on several demographic variables of each county. The coefficients from this regression should be thought of as the impacts of these demographics on the transmission rate. The results from this regression are reported in Table 2. We observe that the contagion in the disease is increased with greater population density and the percentage of commuters who use public transportation. We also observe that contagion rates are higher in areas with a higher fraction of Black and Hispanic residents. Furthermore, the rate of spread is higher for seniors than for younger people, but children and non-senior adults do not seem to have statistically significantly different rates of contagion.

We next measure the out-of-sample prediction accuracy of our model using a hold-out sample of 75 days (May 24 - August 6) to see how well our model forecasts new cases. We use the observed county level of daily social distancing for our out-of-sample predictions. Nationally, this reflects an approximately 50%-60% return-to-normalcy, but this varies quite a bit across the country. We define the percentage return-to-normalcy as 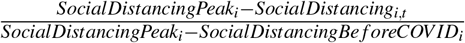, where *SocialDistancingPeak*_*i*_ is the social distancing level in county *i* at its peak (April 5-April 11, 2020), *SocialDistancingBe f oreCOV ID*_*i*_ is the observed lowest level of social distancing in February, and *SocialDistancing*_*i,t*_ represents social distancing level on date *t*. For example, a 25% towards normalcy represents social distancing at the level of 0.25 × (minimum social distancing) + 0.75 × (maximum social distancing).

Figure 1 shows the US actual cumulative cases along with out-of-sample forecasts from a model with *ω* = 0.57 and a standard model with *ω* = 1. The black hashed line represents the actual cumulative cases in the US. The green solid line and the red dashed-line show the out-of-sample forecasts with *ω* = 0.57 and *ω* = 1, respectively. We readily observe that the model with *ω* = 0.57 fits the data well while the model with *ω* = 1 does not. Three states that had their Shelter-in-Place orders expire or stuck down early are Florida, Georgia, and Wisconsin. To further evaluate our model’s accuracy in prediction, we repeat the same out-of-sample prediction comparisons for these three states in Figure 2. The figure again shows that the model with *ω* = 0.57 has a much better fit than the model with *ω* = 1.

**Figure 1.**
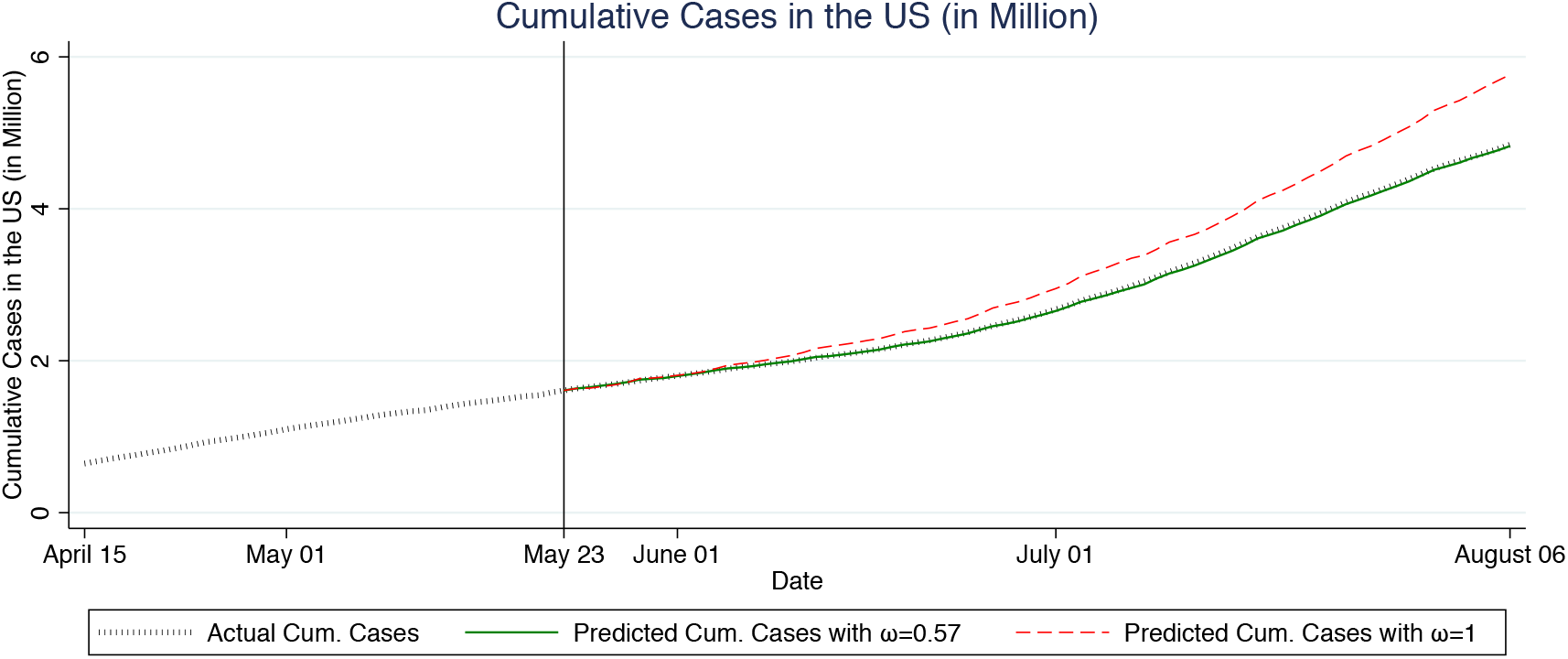
Out-of-Sample Fit Comparisons of the US between Our Model and Standard SIR model. The vertical line on May 23, 2020 indicates the last date used to estimate each model.

**Figure 2.**
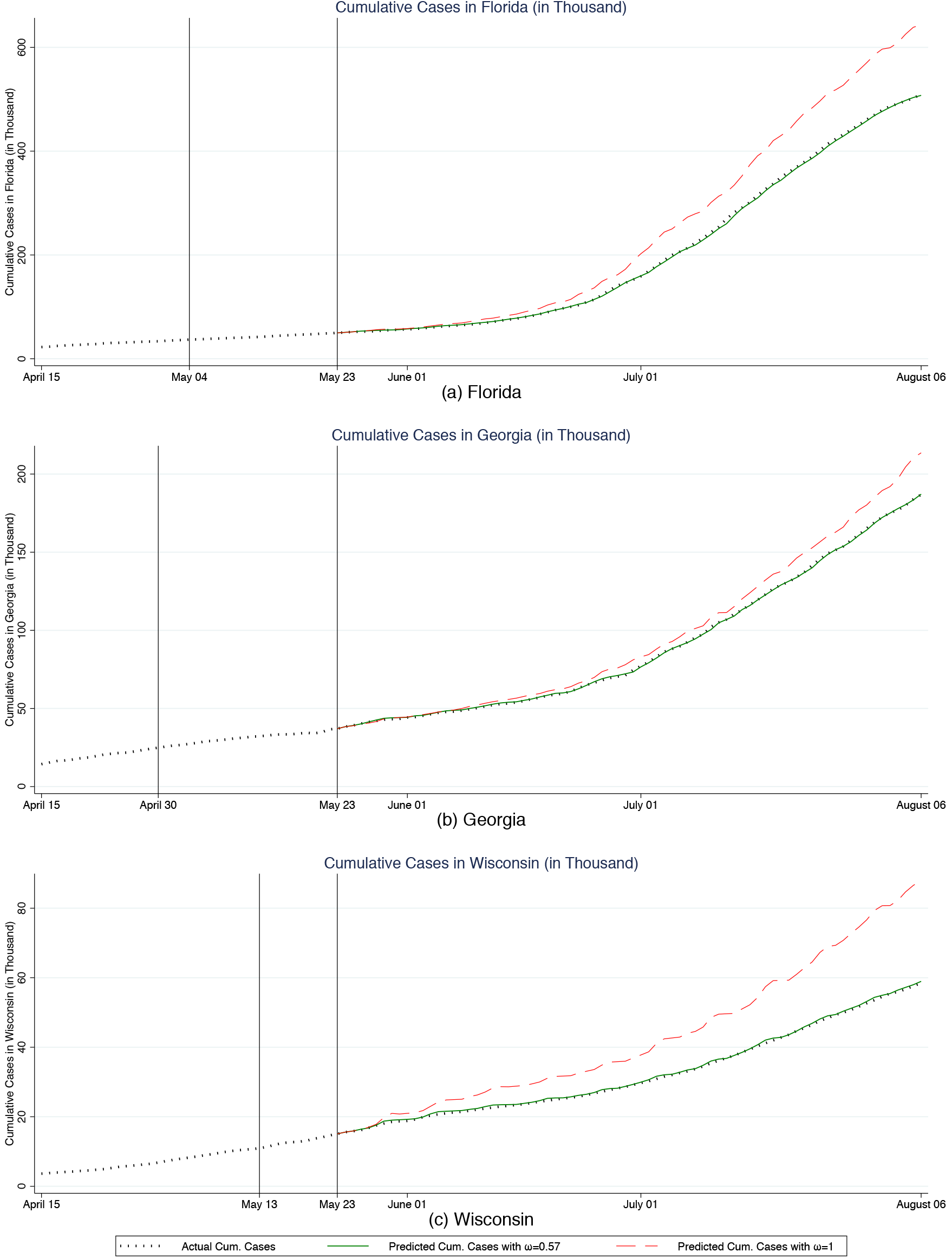
Out-of-Sample Fit Comparisons of Florida, Georgia, and Wisconsin between Our Model and Standard SIR model. The vertical line on May 23, 2020 indicates the last date used to estimate each model. The vertical lines to the left indicate the expiration date of Shelter-in-Place order in each state.

We next simulate daily and cumulative cases from August 7 to October 31, 2020 under different levels of social distancing. When forecasting future cases, we use previous 5-year county temperature data and the May 2020 county average humidity. The top of Figure 3 shows three sets of forecast daily cases after August 6, corresponding to 75%, current, and 25% levels of return-to-normalcy. We observe that social distancing at the current 60% return-to-normalcy first leads to a slightly increasing but then slowly decreasing number of cases, going from around 55,000 cases per day in early August to 25,000 cases per day in the end of October. If the US practiced social distancing at the level reflecting a 25% return-to-normalcy for even a few weeks, new cases would drop to a much lower level of around 9,000 per day. On the other hand, a return to a 75% level of the normalcy would cause cases to surge for about two months. However, after two months cases would again reach a long-term plateau, although this would occur at a level that was almost double of what would be experienced under the early-August level of social distancing. The bottom of Figure 3 depicts the corresponding cumulative cases for the same time period under 100%, 75%, current, 25%, and 0% levels of return-to-normalcy. The figure shows a consistent pattern where the cumulative cases look almost linear after the initial take-offs. There would be substantially more cases if we returned to the pre-COVID level of social distancing.

**Figure 3.**
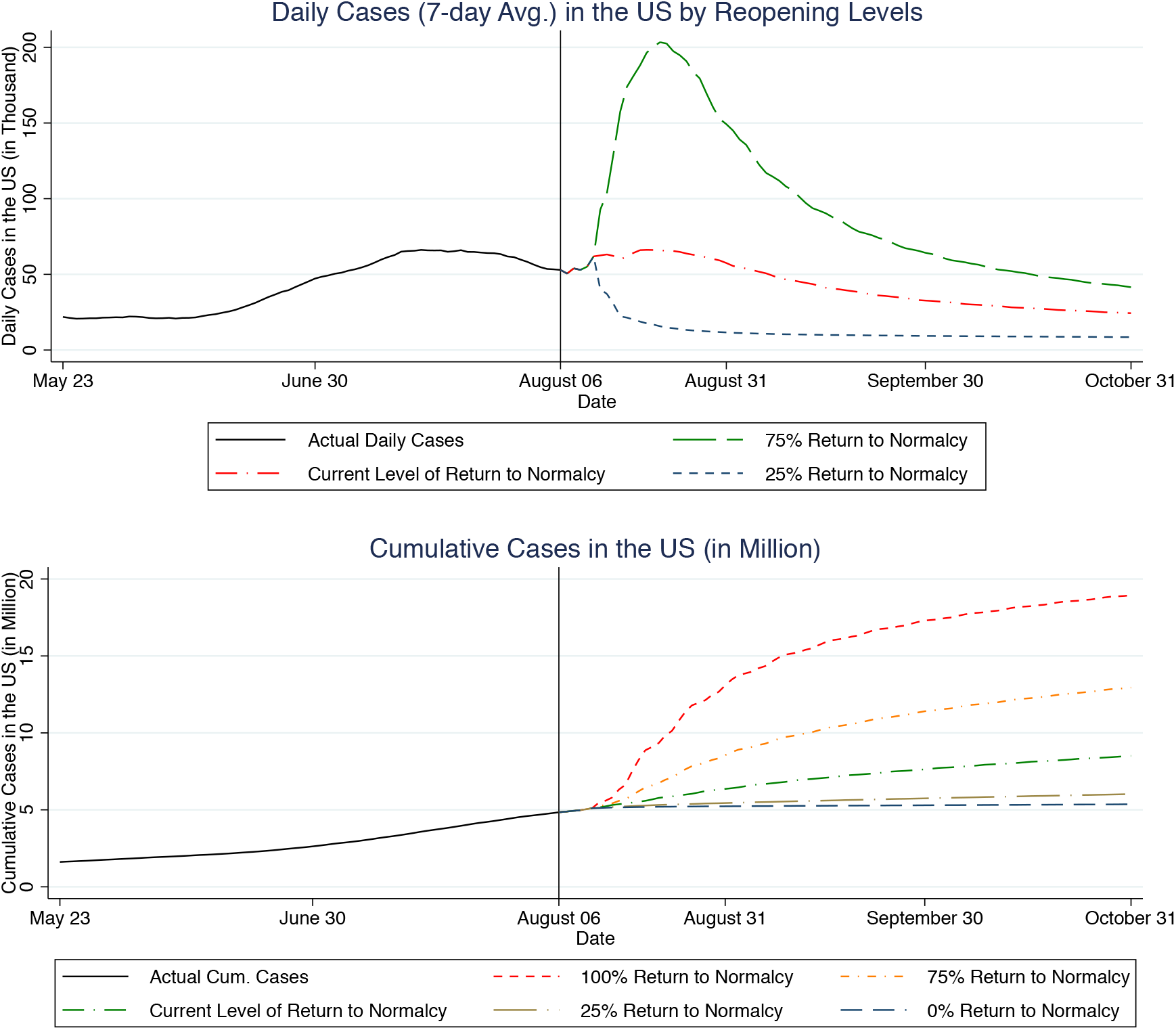
Daily and Cumulative Cases Forecasting under Different Reopening Strategies. The vertical line indicates the last date of case data sample.

## Methods

In this subsection, we detail the assumptions we make and the estimation procedure. The model is laid out in equations 1 and 2 above. For simplicity, we rewrite equation 2 as 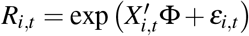, where *X*_*i,t*_ includes county dummy variables, date dummy variables, the measure of social distancing *d*_*i,t*_, and daily average temperature *m*_*i,t*_ and humidity *h*_*i,t*_. Φ is the vector containing the parameters *α, β, λ, µ*, and *θ*, which measures the impact of each element in the vector *X*_*i,t*_ on the transmission rate *R*_*i,t*_. We assume that the errors *e*_*i,t*_ are uncorrelated across counties. We further assume that *e*_*i,t*_ is uncorrelated across time, although we cluster the standard errors by county.

We estimate the model by taking logarithm of both sides. After rearranging we get:

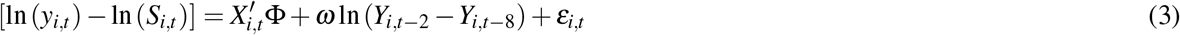

Note that sometimes *y*_*i*.*t*_, the diagnosed case number, is 0 for some counties on some dates. Therefore, we adjust this formula slightly by adding 1 to *y*_*i,t*_ so the logarithmic values are always well-defined:

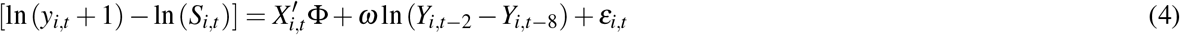

In some counties, *Y*_*i,t-*2_ *Y*_*i,t-*8_ is 0 for some periods. We do not use those observations for estimation. Note that because this is a lagged variable, this is a selection based on independent variables and not based on dependent variables, and hence it does not bias our estimation.

One concern that can arise in estimating this model is that social distancing levels (and regulations) are not determined in a vacuum: Rather, people social distance more in areas that are hit harder by COVID-19. Thus, *e*_*i,t*_ may be correlated with social distancing, causing a biased measurement of the impact of social distancing on the rate of contagion. We thus use an instrumental variables (IV) technique to control for this endogeneity bias, where the amount of rain is our instrument for social distancing. Specifically, we assume that rain directly shifts the level of social distancing, but is not correlated with *e*_*i,t*_ conditional on the temperature and humidity. Several other papers have used rain as an instrument for social distancing^10,11,12,13^. The first-stage *F*-statistic for the strength of rain as an instrument is 214.44, which is highly significant, indicating that rain is a strong instrument.

## Data

Our data come from a multitude of sources. We detail the data sources at https://github.com/songyao21/covid_data_depot. There are a few nonstandard issues to note. Our data on COVID-19 cases consists of county-level, officially confirmed daily case data of 2,924 US counties from February 1 to August 6 (with the last 75 days used as a hold-out sample). COVID-19 also has an incubation period of approximately 5 days^14,15^. Because of this lag from infection to diagnosis, we assume that cases reported on a particular date actually measure the COVID-19 infections from 5 days earlier. We also assume that the true number of cases is approximately 10 times the number of diagnosed cases. We get this number by assuming that the Infection Fatality Rate (IFR) is 0.75%^16^. We also assume that any deaths occur 14 days after the confirmed test results. On May 23, 2020, the last day of our estimation case data, there were 92,622 deaths in the US. On May 9, 2020, there were 1,304,726 officially diagnosed cases. We hence obtain the factor as (92,622/0.0075)/1,304,726 = 9.5. We round this number up to 10. This is consistent with Centers for Disease Control and Prevention (CDC) director Robert Redfield’s estimate of the ratio between actual and confirmed cases^17^. Our estimates are not sensitive to the specific factor we use. When we run the simulations, we divide our model’s predicted case numbers by 10, which gives us the prediction of diagnosed cases.

## Conclusion

We use a modified SIR model to study the impacts of different factors on the spread of COVID-19. We find that the impact of each additional infectious individual decreases as more people become infected. A potential mechanism underpinning this finding is that infections are more likely to occur within interconnected networks. Understanding the shape of this relationship, and the nonlinear aspects of it, are important for understanding how COVID-19 spreads. Unlike previously-estimated SIR models, our model allows for the possibility that the contagion process will grow or shrink at relatively steady levels, whereas traditional SIR models have contagion either taking off exponentially (if *R >* 1) or falling quickly (if *R <* 1).

We further find that social distancing helps to curb the speed of the spread. Consequently, we need to be cautious of breakouts in networks and maintain a reasonably high level of social distancing during the reopening of the economy. Taking the network effects and social distancing effects together helps give more accurate forecasts about the timeline of the disease spread, and the ability to analyze and set policies about when to instate shelter-in-place restrictions or when to allow businesses to be open.

## Data Availability

Most of the data we use is publicly available from multiple sources, and we lay out our sources ad where researchers can access this data clearly. Cell phone stay-at-home rates are from SafeGraph, which is available for free from all researchers studying COVID-19. We cannot share this data, but it is widely available, so the paper is auditable.

https://github.com/songyao21/covid_data_depot

## Acknowledgements

We thank SafeGraph Inc. for providing the mobility data used in our analysis.

## Author contributions statement

All authors formulated the model, crafted the data analysis strategy, and drafted and revised the manuscript. M.L. and S.Y. conducted the data analysis.

## Technical Appendix: Forecasting the Spread of COVID-19 under Different Reopening Strategies*

**Meng Liu Raphael Thomadsen Song Yao**

Olin Business School

Washington University in St. Louis

This draft: August 10, 2020

In this online appendix, we first present our data. We then discuss the sensitivity of our results to the assumed contagious period. Next, we provide some supplemental details to our simulations. Finally, we lay out how the concavity we estimate could come from a model of interconnected networks.

### A Data

Our data come from a multitude of sources. We lay out the sources for each of these in turn.

#### A.1 Positive Cases

Data of positive cases are based on the COVID-19 data published by the New York Times (https://github.com/nytimes/covid-19-data, accessed on August 6, 2020). The data contain the daily confirmed case counts for 2,953 U.S. counties or county-equivalents. The case data for the five boroughs of New York City, however, are not recorded separately by New York Times. In this case, we use the data published by the Health Department of New York City in lieu of the five boroughs (https://github.com/nychealth/coronavirus-data, accessed on August 6, 2020). The case data of Kansas City, Missouri are also recorded separately because it overlaps with 4 adjacent counties. We attribute the cases of Kansas City to Jackson County, Missouri because most of the city lies within Jackson County. We also drop 3 counties because we do not have social distancing data for 2 of them, and we cannot match the population data for a third (Oglala Lakota County, SD). Finally, we remove counties that had no confirmed cases during our estimation sample period of Feb 1, 2020 to May 23, 2020. After all the above-mentioned filters, we have a panel of 2,924 counties. These counties account for 99.76% of the US population and 99.91% of the total U.S. confirmed COVID-19 cases till August 6, 2020.

There are a few days where there are negative cases that are reported. These are generally corrections to previous over-reporting. Thus, we clean the negative numbers of cases by subtracting the absolute value of the negative cases from the proceeding day. In the event that that leads to a negative number of the proceeding day, we iterate again.

#### A.2 Social Distancing

We use social distancing data from the company SafeGraph, which collects cellphone GPS data from U.S. residents, and has made them available for free to academics studying COVID-19. These data are collected through a series of pings that the company receives for all users who have installed a number of smartphone apps. The list of apps that collect this information is kept as a trade secret. We measure social distancing as the first principle component of several measures. The measures are, on a given day, percentage of residents staying home, percentage of residents working at workplace full-time, percentage of residents working at workplace part-time, median duration of residents staying home, and median distance of residents traveled. The SafeGraph data are published at the Census Block Group level. To accommodate other data sources which are available at a less granular level, we aggregate the this variable to the county level by taking the weighted median, using the number of cellphones in each Census Block Group as the weight.

#### A.3 Demographic data

We obtain the demographic data from the Census Bureau’s 2014-2018 American Community Survey (ACS), which contains information of each county’s profile of population, ethnicity, age, median income, and commuting pattern. The ACS, however, does not report population densities. Safe-Graph, the company who provides us with the social distancing data, also maintains a dataset of the land area of each Census Block Group in the US. We aggregate the land areas to the county level. Together with the county population information from the Census Bureau, we are able to construct the population density data of each county.

#### A.4 Weather data

We gathered historical daily rain and temperature data from National Oceanic and Atmospheric Administration (NOAA) (source: https://www.ncei.noaa.gov/metadata/geoportal/rest/metadata/item/gov.noaa.ncdc:C00861/html, accessed on May 21, 2020). The raw weather data is at the weather station level and we match weather stations to the counties they are in. We use the average values across weather stations within the same county to construct the weather variables for that county. For a small number of counties where there are no associated weather stations, we use the daily state averages as proxies.

#### A.5 Putting it all together

Our sample is an unbalanced panel because counties start to have positive number of confirmed cases on different dates. The earliest date we observe in the sample is Jan 29, 2020, and the last day is August 6, 2020. Note that we construct actual cases using reported cases 5 days later, and thus the corresponding sample period based on reported cases is Feb 3, 2020 to August 1, 2020.

Summary statistics of all of the variables we use in the estimation are presented in Table A1. Note that our humidity data end on May 18, 2020. For estimation we only use case data up to May 18. But our case data proceed past the dates used for estimating the model and we use those data for validating the model. Those data are publicly available, and we have also posted the compiled case data and the code for compiling the data at https://github.com/songyao21/covid_data_depot.

### B Sensitivity to Duration of Contagious Period

Research on COVID-19 is nascent, and there are different views of how long infected individuals stay contagious. Suppose that such individuals are contagious for 14 days instead of 6 days. Then the model becomes:

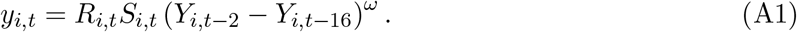

We present the estimation results of this model in column 2 of Table A2. Note that this regression has more observations because there are fewer instances where we observe no cases in a county for a 14-day window than for a 6-day window. The results are largely unchanged. The coefficient on social distancing levels are slightly lower, but well within one standard error of the corresponding coefficient in column 1. The exponent on the infectious individuals is 0.523. That is somewhat smaller (but statistically different) than the 0.571 we observe with the shorter 6-day window, but overall the curvature shape is similar to what we have observed with the 6-day window. As we will discussion below in Section C, both specifications give similar long-run forecasting results.

**Table A1:** Summary Statistics.

|  | (1) | (2) | (3) | (4) | (5) |
| --- | --- | --- | --- | --- | --- |
| Time-varying Variables | N | Mean | Std Dev. | Min | Max |
| Reported cumulative cases | 131,272 | 337.4 | 2,178 | 0 | 63,690 |
| Reported new daily cases | 131,272 | 11.40 | 64.40 | 0 | 2,174 |
| Average temp (in Celsius) | 131,272 | 12.65 | 6.733 | -16.40 | 32.06 |
| Rain (mm) | 131,272 | 3.469 | 7.937 | 0 | 145.1 |
| Humidity (percentage) | 131,272 | 66.95 | 15.24 | 8.745 | 100 |
| Social_distancing | 131,272 | 1.115 | 1.029 | -4.578 | 4.878 |
| Time-invariant Variables |  |  |  |  |  |
| Median income (\$) | 2,923 | 52,580 | 14,606 | 19,391 | 144,821 |
| Population | 2,924 | 111,906 | 344,510 | 625 | 1.004e+07 |
| Population density (people / square miles) | 2,924 | 292.5 | 1,860 | 0.0359 | 71,891 |
| Share of population senior ($\geq 70$) | 2,924 | 0.122 | 0.0327 | 0.0229 | 0.384 |
| Share of population youth ($\leq 17$) | 2,924 | 0.224 | 0.0338 | 0.0732 | 0.403 |
| Share of population black | 2,924 | 0.096 | 0.148 | 0 | 0.874 |
| Share of population Hispanic | 2,924 | 0.092 | 0.135 | 0 | 0.991 |
| Share of population public transit commuters | 2,923 | 0.010 | 0.0333 | 0 | 0.642 |

We next regress the county fixed effects on several demographic variables, which are reported in Table A3. Comparing the results for the 6 day vs. 14 day contagious periods, we see that over all the results are qualitatively very similar. The main difference is that if the contagious period is 14 days we observe that children are also statistically more contagious than non-senior adults.

### C Simulation

We forecast the cumulative and daily cases of COVID-19 through the end of October at different levels of social distancing. Those forecasts appear in Figure 2 in the original paper. In Figure A1 below, we replicate the graph with cumulative cases, but further add confidence intervals. To avoid cluttering, we only depict current and 75% return-to-normalcy levels in A1. We observe that these forecasts are indeed statistically significantly different.

As a robustness check regarding the 6-day contagion window specification, we also consider forecasting US daily cases under the specification where the contagion window is 14 days. Figure A2 shows the evolution of daily cases till October 31, 2020 under current-level (early August) and 75% return-to-normalcy regimes. We overlay the forecasts of both 14-day and 6-day specifications for easy comparison. From the figure, we may see the forecasts of 14-day and 6-day contagion window specifications are fairly close.

**Table A2:**
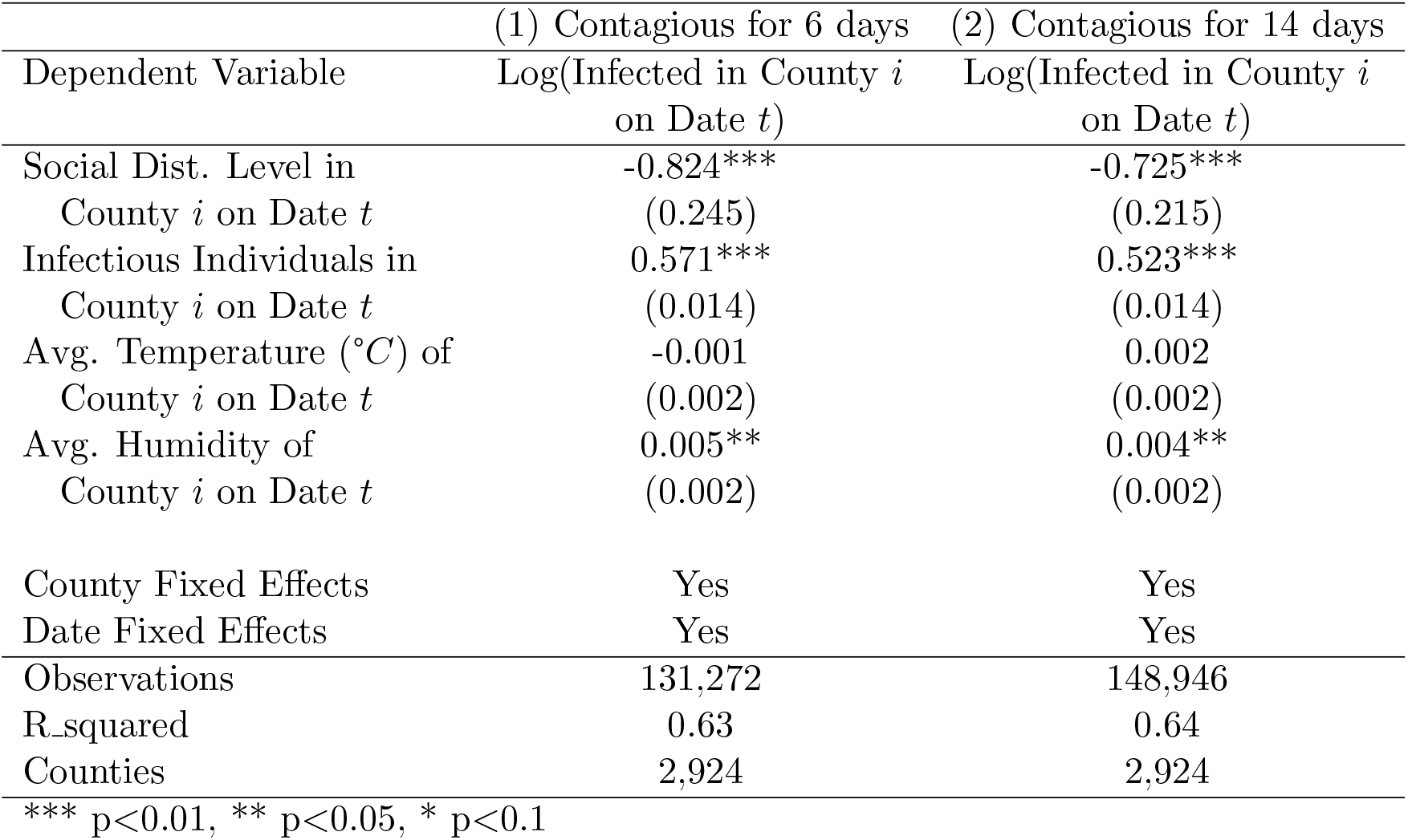
Estimation of a Modified SIR Model.

|  | (1) Contagious for 6 days | (2) Contagious for 14 days |
| --- | --- | --- |
| Dependent Variable | Log(Infected in County $i$<br>on Date $t$) | Log(Infected in County $i$<br>on Date $t$) |
| Social Dist. Level in<br>County $i$ on Date $t$ | -0.824***<br>(0.245) | -0.725***<br>(0.215) |
| Infectious Individuals in<br>County $i$ on Date $t$ | 0.571***<br>(0.014) | 0.523***<br>(0.014) |
| Avg. Temperature ($^{\circ}C$) of<br>County $i$ on Date $t$ | -0.001<br>(0.002) | 0.002<br>(0.002) |
| Avg. Humidity of<br>County $i$ on Date $t$ | 0.005**<br>(0.002) | 0.004**<br>(0.002) |
| County Fixed Effects | Yes | Yes |
| Date Fixed Effects | Yes | Yes |
| Observations | 131,272 | 148,946 |
| R_squared | 0.63 | 0.64 |
| Counties | 2,924 | 2,924 |

\*\*\* $p < 0.01$ , \*\* $p < 0.05$ , \* $p < 0.1$

### D Concavity of SIR model and Network Dynamics

A unique feature of our model is that we estimate an exponent on the number of contagious cases. We include this flexibility because such a model fits the data much better, and also leads to forecasts that have more limited growth after an initial take-off of COVID-19 cases, as is commonly observed. We illustrate that the concave relationship we estimate for the number of contagious individuals on the number of new cases can come from social networks between people through a very simplified model of networks and disease process.

To do this, we simulate a network with the following process: We take 10,000 individuals. We create a network by first randomly assigning that any two individuals will be joined with a common node with probability 11/20,000. Call these connections “round-1 friends.” We then expand this network by assigning each node to have an edge with each of the friends of round-1 friends with a probability of 0.8.

We assume that the disease spreads with the following process. We seed 4 individuals to have the disease in period 0. Then in each period we assume that any connected individual will get sick with probability 0.4.

After simulating this process, we then regress (ln (*y*_*t*_) *-* ln (*S*_*t*_)) = *c* + ω ln (*y*_*t-*1_)+ ε_*t*_. The mean value for 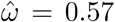. This shows the plausibility of network effects leading to an estimate in the range that we have estimated in our main model. We have placed the R code for this simulation at https://github.com/songyao21/covid_data_depot/network_simulation, so that interested readers can play with the parameters to understand the process more.

**Table A3:** Analysis of County Fixed Effects.

|  | (1) Contagious for 6 days | (2) Contagious for 14 days |
| --- | --- | --- |
| Dependent Variable | County Fixed Effect | County Fixed Effect |
| Log(Pop. Density)<br>(People/Sq. Miles) | 0.4410***<br>(0.0096) | 0.4006***<br>(0.0095) |
| Fraction Black Residents | 0.8084***<br>(0.0979) | 0.7658***<br>(0.0970) |
| Percentage Hispanic<br>Residents | 1.3438***<br>(0.1003) | 1.3112***<br>(0.0993) |
| Percentage of Commuters<br>using Pub. Transportation | 5.0215***<br>(0.4140) | 5.5818***<br>(0.4101) |
| Log(Median Income)<br>(in U.S. dollars) | 1.3387***<br>(0.0579) | 1.2564***<br>(0.0573) |
| Percentage of Senior<br>Residents ($\geq 70$ yrs) | 1.8825***<br>(0.5255) | 2.2584***<br>(0.5205) |
| Percentage of Children<br>Residents ($< 18$ yrs) | 0.6614<br>(0.4733) | 1.0609**<br>(0.4688) |
| Constant | -16.6781***<br>(0.6462) | -15.7657***<br>(0.6401) |
| R_squared | 0.69 | 0.66 |
| Counties | 2,923 | 2,923 |

\*\*\* p<0.01, \*\* p<0.05, \* p<0.1

**Figure A1:**
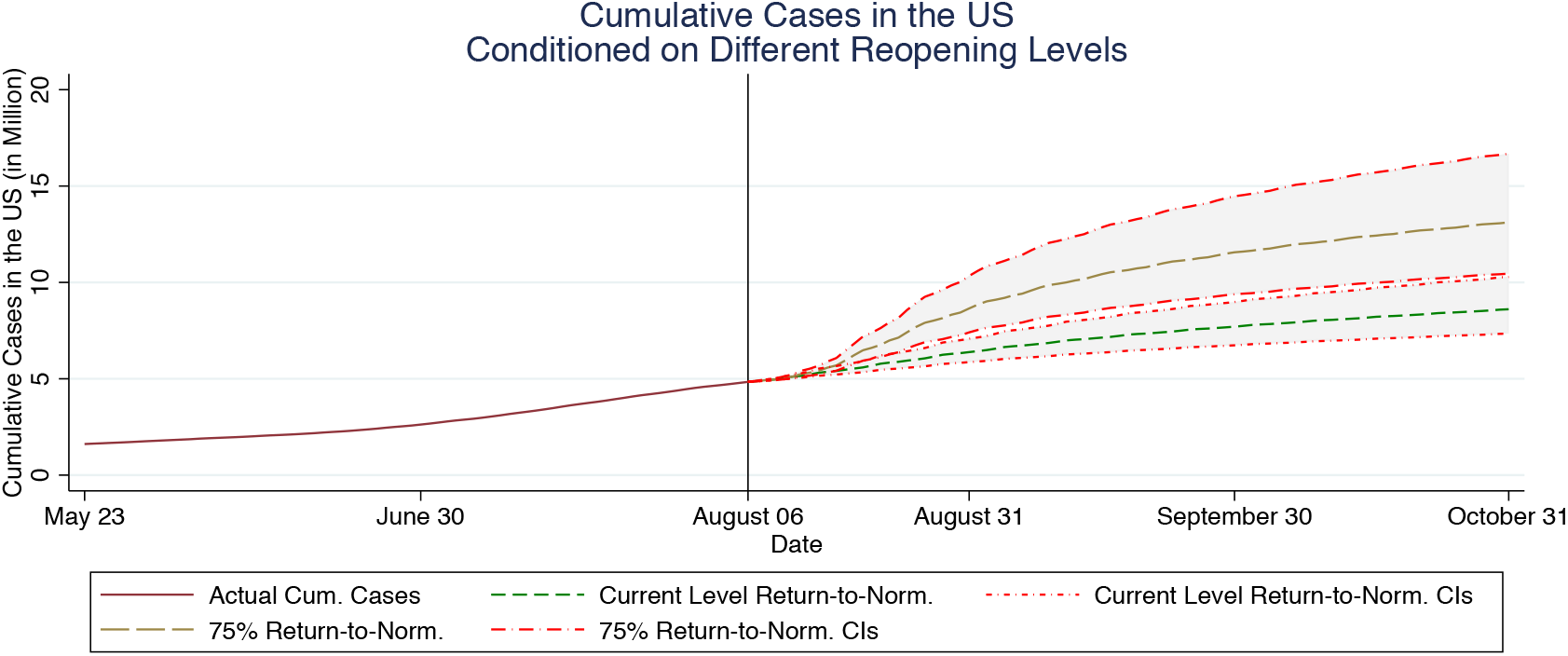
Cumulative Case Forecasting under Different Reopening Strategies with Confidence Intervals. The vertical line indicates the last day of diagnosed case data sample.

**Figure A2:**
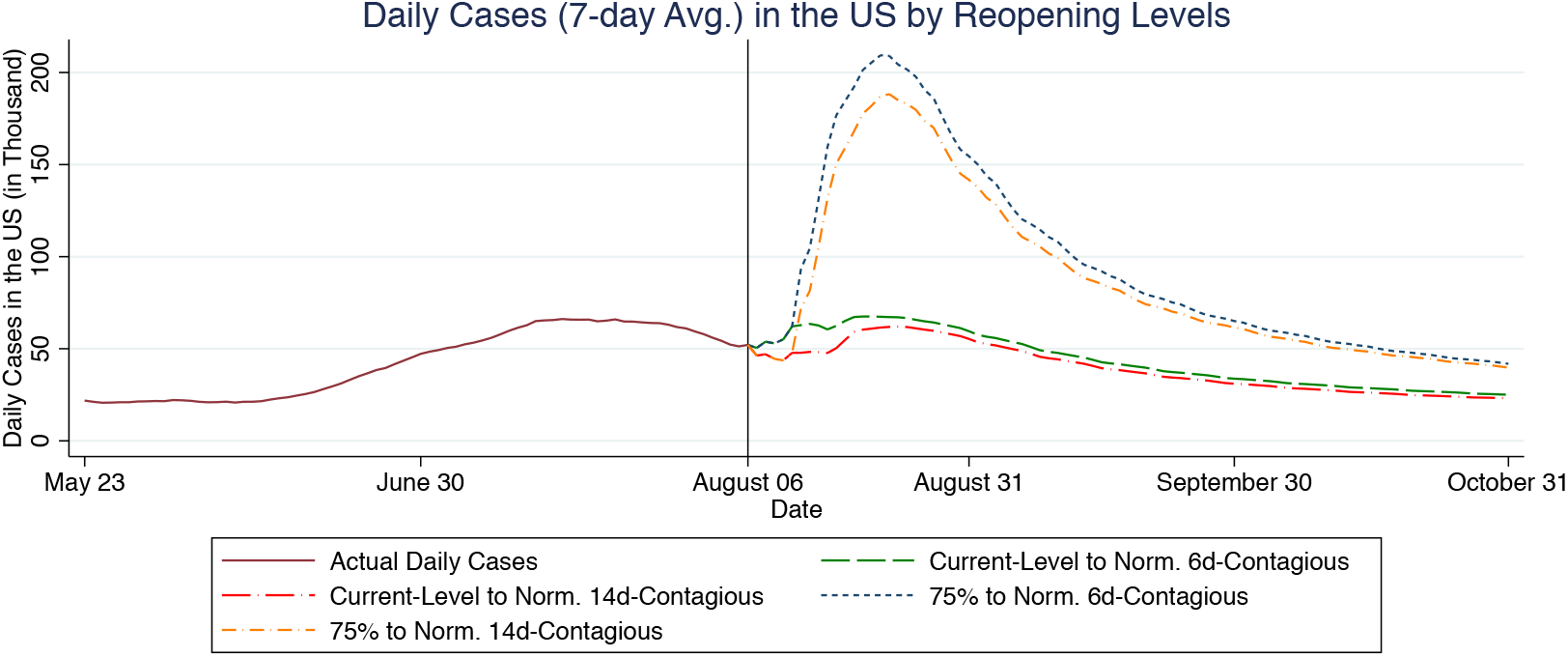
Daily Case Forecasting under Different Contagion Window Specifications. The vertical line indicates the last day of diagnosed case data sample.

### Footnotes

* Please contact Liu, Thomadsen, or Yao for correspondence.

